# Expert perspectives on improving services for patients with periprosthetic femoral fractures: a qualitative study

**DOI:** 10.64898/2026.07.01.26357068

**Authors:** Helen Gibson, Choon Key Chekar, Dawn Goodwin, Cliff Shelton, Toby O Smith, Antony Johansen, Mohammad Aryaie, Walter Muruet, Mike Reed, Jonathan T Evans, R Whitehouse Michael, Mark Baxter, Alex Bottle, Jonathan Benn

## Abstract

**Background:** The incidence of post-operative periprosthetic femoral fractures (POPFFs) is increasing. However, specific clinical guidance relating to patient management does not exist, resulting in variations in care and outcomes. This study aimed to elicit and synthesise expert knowledge in POPFF service delivery and explore views on variations in service provision and the factors influencing these.

**Methods:** Semi-structured interviews were undertaken with healthcare professionals with expertise in POPFF care from England and Wales to explore current practices, challenges, service variations and perceived future opportunities. Participants were identified through specialist research and clinical networks for POPFF and hip fracture care, authors of key publications on the subject, national leads for POPFF/hip fracture networks, and research team contacts. Interviews were analysed using thematic analysis.

**Results:** Ten interviews were undertaken with experts in POPFF services across a range of professional roles. Four themes were identified: conceptualisation of POPFF (by different professional groups and in different service settings) and understanding of POPFF patient needs; sources of variation in management and care of POPFF patients; service model rationales, advantages and disadvantages; and potential strategies to improve POPFF care.

**Conclusion:** When designing POPFF services, we suggest that four key areas need consideration: the extent to which POPFF patients are a distinct group with particular care needs; the necessity for and consequences of patient transfer between wards and hospitals; the resourcing of extensive multidisciplinary support for POPFF patients; and the need for national initiatives to encourage service developments.

These findings should form the basis of future clinical guidance. Sensitivity to contextual factors driving variation in services is needed to ultimately improve care for POPFF patients.

## Introduction

The incidence of post operative periprosthetic femoral fractures (POPFFs) is increasing and is linked to the rising number of hip and knee arthroplasties and increased life expectancy of patients with a prosthesis. Between April 2015 and December 2018 there was a 13% year-on-year increase in admissions for periprosthetic fracture in England (1). The incidence of POPFF is generally higher in women and those aged over 60 (2). Treatment costs are high (3), and patients often have multiple comorbidities and experience long hospital stays (2).

In the UK, primary hip fractures are managed with reference to specific multiprofessional clinical guidance (4,5). However, similar guidance for POPFF does not exist. As such, POPFFs are not subject to a best practice tariff (payment available to NHS England Trusts to incentivise treatment according to guidance), nor are they part of the NHS England’s Get It Right First Time (GIRFT) initiative, which aims to reduce national practice variations. In common with primary hip fractures, however, POPFFs are now monitored by the National Hip Fracture Database (NHFD), which captures information on patients in England, Wales and Northern Ireland with the aim of delivering improvements in care. In 2020, the NHFD began to capture data on POPFFs, and in 2022, the KPIs used by the NHFD were applied to POPFFs. However, because of differences in the patient characteristics and approaches to management, there is some uncertainty about the transferability of hip fracture KPIs to patients with POPFF (6). Insights for POPFF care may be gleaned from the organisational and service delivery factors known to impact outcomes for hip fracture patients. For example, for patients with hip fracture, assessment by an orthogeriatrician within the first 72 hours of admission was associated with cost savings of £529 per patient and a lower 365-day mortality rate (7).

Whilst there has been an increase in epidemiological data on POPFF and growing evidence of differences in management and outcomes (8), there remains a paucity of qualitative research on the factors driving variations in the design and delivery of POPFF care. There is also an increased awareness of the need for greater incorporation of qualitative designs into orthopaedic research and practice (9). By generating rich, detailed accounts, qualitative research can help address the ‘how’ and ‘why’ of a phenomenon thus complementing quantitative research. This study aimed to elicit and synthesise expert knowledge of POPFF service delivery to address the following questions:

1. What are the challenges associated with POPFF services and patients, compared with hip fracture care?
2. What are the established service delivery models for POPFF care and how do they vary?
3. What are the service design considerations and organisational factors influencing variations in POPFF care and service design?
4. What are the opportunities for enhancement of the design and delivery of services for POPFF patients?

## Methods

### Design

Semi-structured interviews explored expert perspectives on variations in POPFF service provision, including contextual factors driving variations and the rationales underpinning specific service configurations. This qualitative study is part of the mixed-methods PROFOUND study (Periprosthetic Femoral Fractures Outcomes, Management and Data – see Acknowledgments). It was approved by Leeds University School of Psychology Research Ethics Committee (reference: PSCETHS – 722).

### Participants and recruitment

A purposive sampling strategy was used to recruit professional and policy representatives with expertise in POPFF care from England and Wales. Potential participants were identified through multiple channels, including specialist research and clinical networks for POPFF and hip fracture care, authors of key publications on the subject, national leads for POPFF/hip fracture networks, and research team contacts and were invited to participate via email. All contacted experts agreed to participate. Whilst our interviewees were selected to represent varied professional and regional perspectives on the design of POPFF services, we did not aim to systematically sample all possible regions or service design variations. However, to ensure that the views expressed did not emphasise any single type of service or perspective, we: 1) included experts in centralised roles associated with national audit and policy (i.e. with experience across multiple service configurations), 2) asked experts to describe local services and comment on service variations in the generic sense, and 3) used specific follow-up questions designed to probe for counter examples or the extent to which a feature was common across localities. Ten interviews were undertaken with healthcare professionals from a range of backgrounds representing the various components of POPFF care services.

### Data Collection

Data collection was conducted between January 2024 and May 2024. An interview topic guide (Table 1) was developed based on the PROFOUND study research questions ([The PROFOUND study | Faculty of Medicine | Imperial College London](https://www.imperial.ac.uk/school-public-health/primary-care-and-public-health/research/the-profound-study/)), review of relevant literature (10–13) and feedback from the multidisciplinary research team. It covered three main areas: quality of care for POPFF; service delivery models; and the impact of variations in service delivery. The interview topic guide was piloted with one participant and refined accordingly. All interviews were conducted virtually using Microsoft Teams by three members of the research team (DG, CKC, JB). All three interviewers are experienced qualitative researchers with sociological and psychological expertise in healthcare organisation, work and safety, and one researcher (DG) also has a background in nursing. To enhance the internal consistency of the interview format, initial interviews were conducted by pairs of researchers, and to reduce researcher subjectivity multiple iterations of coding were undertaken drawing on the skills and expertise of each of the researchers.

**Table 1.** Interview topic guide Topic.

| Topic | Example questions |
| --- | --- |
| Quality of care for POPFF patients | What is the experience of POPFF like for patients?<br>Why should we focus on improving services for this group?<br>What factors do you think are critical to good patient outcomes for POPFF?<br>What service design/delivery factors are important for effective POPFF care?<br>How should we be measuring quality of POPFF care in existing services? |
| Service delivery models | What are the most common service delivery models for POPFF patients?<br>Can you describe what typically happens in the care of POPFF patients under these models and how they differ?<br>What is the rationale for why services are designed in this way?<br>Are you aware of any variations in service delivery models for POPFF and in what ways they can vary? |
|  | What drives variation in service models? E.g. regional context, patient population, etc |
| Impact of variations in service delivery | How would you evaluate [service model X e.g. hub & spoke model/self-sufficient model]?<br>What are the advantages and disadvantages of [approach X]?<br>Do you think that having variable service models is useful/efficient/effective? Why?<br>What is the impact on quality of immediate/long-term care/QoL/patient safety?<br>What model or approach would you suggest for service configuration at local/national level and why? |

Interviews lasted up to 60 minutes, and were transcribed verbatim by a professional transcription service, external to the University.

### Data Analysis

Interviews were analysed by a multidisciplinary research team using thematic analysis and utilized the six steps described by Braun and Clarke (2006) (Table 2)(14). Illustrative quotes are used in this paper to present key concepts from the analysis. Throughout the process, qualitative data analysis software (NVivo, Lumivero, Denver, USA) was used to assist with organising the data and facilitate the coding and development of themes. Initial codes were developed by one researcher with a background in social sciences and refined with feedback from the broader multidisciplinary team at research group meetings, including input from clinicians in relevant fields and lay members (former POPFF patient). Subsequent coding was completed iteratively with input from three principal researchers with expertise in psychology, healthcare quality and safety and health service research (step 2). The coding framework was then applied to each interview transcript by one researcher and to improve reliability and validity two researchers each coded two transcripts independently. Codes were then collated into themes which included example quotes and were shared for refinement and feedback in the qualitative research team meetings. (steps 3,4,5).

**Table 2.** Application of Braun and Clarke’s (2006) thematic analysis.

| **Steps of thematic analysis** |  |
| --- | --- |
| 1. familiarisation with the data | Three qualitative researchers with expertise in psychology, medical sociology, healthcare quality and safety read the interview transcripts. |
| 2. generating initial codes | They then independently coded the first two transcripts drawing on literature as well as working inductively to ensure codes accounted for all the data. The initial coding framework was collectively reviewed by the three researchers to establish reliability and validity of coding and a further 3 transcripts were coded. |
| 3. searching for themes | Candidate themes and definitions were developed through review and discussion of the initial coding framework by the qualitative research team (consisting of the three researchers about plus a clinician with expertise in qualitative methods). |
| 4. reviewing themes | Themes were reviewed and discussed by the broader multidisciplinary research team including input from clinicians in relevant fields and lay members (former POPFF patient and spouse). |
| 5. defining and naming themes | Following multidisciplinary discussion with the broader research team, the initial themes were refined and expanded. Once finalised, the emergent coding framework was subsequently applied by a further qualitative researcher across the whole dataset for consistency in coding. |
| 6. producing the report | The final draft of the analysis was reviewed by the broader multidisciplinary team which, as with the initial coding, included feedback from |
|  | clinicians, subject matter experts and lay members. |

## Results

Ten interviews were undertaken with experts in POPFF services across a range of clinical roles and geographic location of services (anaesthesia *n*=1; orthopaedic and trauma surgery *n*=4; orthogeriatrics *n*=2; physiotherapy *n*=1; pre-hospital trauma care *n*=1; trauma nursing *n*=1).

Four themes were identified:

### Theme 1: Conceptualisation of POPFF and understanding of POPFF patient needs

Variations in how POPFF was conceptualised by different professional groups and in different service settings were evident. Some professionals viewed POPFF primarily as an orthopaedic condition while others viewed it as a geriatric condition or a subspeciality of geriatrics.

> *I think hip fractures have always been recognised as a frailty presentation for at least the last 20, 20-odd years, so they’ve always been incorporated as part of geriatric practice […] slowly but surely there’s been a clear spread that these are part of geriatric work and it’s a subspeciality of geriatrics. I think periprosthetics were often in the past seen as an orthopaedic complication rather than being a frailty presentation and over the last few years there’s been a recognition the patient group[s] are very similar.* (Interviewee 2, Orthogeriatrics)

In general, conceptualising POPFF as an ‘entity’ itself, as distinct from hip fracture patients, was perceived as relatively unusual. Some interviewees also viewed POPFF patients as similar enough to hip fracture patients so as to benefit from the same clinical pathway.

> *I’m pretty binary on that. […] If you put the patient at the centre of it, I can’t see how there will be a benefit from somebody not being treated in an established pathway.* (Interviewee 10, Orthopaedic and Trauma Surgery)

Conversely, whilst some interviewees suggested that POPFF patients overlapped with hip fracture patients in their surgical and care needs, they also had distinct differences in that POPFF surgery was more extensive than hip fracture surgery, resulting in POPFF patients’ experiencing more pain and being more likely to require blood transfusions. POPFF patients, it was suggested, required more specialist input, specifically from clinicians familiar with the equipment and devices used to treat some types of fracture.

> *[…] a higher risk of pressure sores because of the splints that they have on, and also a lot of the nurses, even on the specialist ward, don’t come across them that often. So, they do need a lot of […] specialist input to look after the frames and to monitor that they’re fitting properly and not causing sores.* (Interviewee 4, Trauma Nursing)

From a physiotherapy perspective, POPFF patients’ needs were distinct from those of hip fracture patients due to many patients having limited weight bearing capacity post-surgery.

> *The big challenge with periprosthetic fractures from a physiotherapist perspective is the reduction or the limitation of weight-bearing, so it’s common […] that patients will be restricted in their weight-bearing, and that can range from toe-touch weight-bearing […] to full weight-bearing* (Interviewee 6, Physiotherapy)

The rehabilitation period for POPFF patients was thought to be typically longer than for hip fracture patients due to a high incidence of frailty combined with complex injury.

> *Because of the nature of the patient, they’re quite often elderly and frail […] there’s a rare few who can go home within a couple of days, more common that they go into an extended rehab type setup within the Trust.* (Interviewee 3, Orthopaedic and Trauma Surgery)

More complex surgery, pain management, blood transfusions, and extended periods of rehabilitation all necessitated the input of multiple specialist resources. As one interviewee commented.

> *It’s the resources that are the real challenge to be able to deliver what we should be delivering for this group of patients.* (Interviewee 6, Physiotherapy)

### Theme 2: Sources of variation in management and care of POPFF patients

The surgical intervention selected for POPFF patients, i.e. revision or fixation, was considered to vary according to a number of contextual centre and service-level factors. In addition to the local surgeon’s preference, local access to specific surgical expertise and availability of theatres were all considered to be factors.

> *You may hang around waiting for the right surgeon, then need a surgical operating list that’s got the right surgeon at the right time to have the right fixation, so you may have a delay for surgery* (Interviewee 2, orthogeriatrics)

Where specialist surgical expertise was required, the impact on waiting time between admission and surgery was something that all interviewees raised.

> *All of that is expert surgery that can’t just be done by any orthopaedic surgeon, but you tend to need to wait for somebody who’s a specialist in either joint revision surgery, or at least lower limb surgery. And so, the patient will then have a variable period of time waiting for a list where one of those surgeons is available.* (Interviewee 1, Anaesthesia)

The type of ward on which the patient was placed also varied, influencing proximity to appropriately skilled staff and thus the opportunity for POPFF patient-centred care. Specialist wards were seen as preferable. However, wider organisational pressures meant this was not always possible.

> *So, if you come into a busy hospital, it’s very tempting for the emergency department to say “we need to get you out of the emergency department into a ward, we’ll send you off there” […] but actually the patients are having a dreadful experience because they’re on a specialist eye ward, they’re on a medical ward, not on the ward where people have an experience of looking after somebody with a broken hip with confusion and problems, or actually experience of sending such a patient to theatre, getting them back from theatre […] So getting to the right ward means […] you also get to a nurse who understands your condition and a therapist who will start talking to you about when you’ll be getting up and an occupational therapist who can start talking to you about what your home’s like and how are we going to get you back home? And a geriatrician who’s working with that group as a team rather than the geriatrician and the surgeon who are visiting somebody else’s ward with nurses and therapists who don’t understand that sort of patient.* (Interviewee 7, Orthogeriatrics)

Size of the NHS trust, and specifically the availability of dedicated trauma theatres, was considered to be important, with larger centres having a higher throughput of POPFF patients, thus enabling the development of more specialist expertise and capability.

> *I think it’s dependent on the experience of the surgeons that they have on the Trust and how often they do the surgeries. For our surgeons, they do them relatively routinely and they are experienced. I think, you know, if you have experienced hip surgeons and they still do trauma lists and they do get the periprosthetic fractures in as routinely as we do, then I’d say it’s a fairly standard service, but if they’re few and far between and they don’t have a specialist hip surgeon then I can imagine that they will need to transfer them elsewhere.* (Interviewee 4, Trauma Nursing)

Access to post-surgery rehabilitation and appropriate facilities was considered variable by some interviewees, with subsequent effects on patient experience and outcome.

> *We restore anatomy with revisions and with fixation but we don’t restore their muscle bulk and function and I think a massive confounder will be access to care and rehabilitation going forward. So you might have a periprosthetic that’s fixed very quickly, fixed very well and then you’ll lie in bed for 3 weeks because you can’t get to rehab and you know, the experience, everything, might be considerably worse.* (Interviewee 2, Orthogeriatrics)

Effective rehabilitation, therefore, must be commenced in a timely manner with appropriate follow-up, if the benefits of prompt surgery are to be realised. Some interviewees considered that variation between POPFF services was more apparent in the post-discharge stage, as opposed to in the acute setting.

> *I think where the variability is is what happens when patients go home […]. After hospital discharge, I think if I’m honest it’s a bit of a lottery and unfortunately this is a case across trauma and orthopaedics in general with rehabilitation […] there’s some units where they seem to have better community rehabilitation networks, so in our region […] for instance […] historically a better community rehabilitation model where patients can get care and rehabilitation either at home or they come in for outpatient rehabilitation, whereas in [other city], for instance, there’s real variability there and there’s some patients that I suspect slip through the net.* (Interviewee 6, Physiotherapy)

### Theme 3: Service model rationales, advantages and disadvantages

Interviewees were asked to describe how POPFF care was organised and delivered in their own locality and to comment on the extent to which variations were common across services. Two broad models – “hub and spoke” and “self-contained” – were described with the distinguishing feature between the two being the extent of patient transfer. In some large, self-contained centres, there were few transfers and patients received treatment at the hospital at which they first presented.

> *So, we’re a busy centre, we don’t routinely take transfers in, primarily because most of our local trauma units can manage PPFFs, so there will be the odd one or two that come in but [locality is] a very big and busy DGH…….* (Interviewee 2, Orthogeriatrics)

In other localities, POPFF patients were often transferred to a different “hub” hospital to which they were first admitted for treatment, though the degree to which this relationship was formalised varied.

> *I think that in [locality] they tend to take [POPFF patients] from the surrounding area as well […] I suppose it’s more a hub and spoke model […] there’s essentially a hub and I think everything is sent in there.* (Interviewee 3, Orthopaedic and Trauma Surgery)

However, there were also examples of care organisation and delivery that did not fit neatly into one of the two aforementioned models.

> *We’re a big teaching hospital but our elective operating and our hip revision surgeons are in a different hospital. […] So we’re not quite a hub nor a spoke. For the majority of people who come in with a periprosthetic fracture they will be able to have surgery done in my hospital. Some of them will get transferred over to the elective part of my hospital, […] but they’ll get transferred over to the other site within the hospital for specialist revision surgery.* (Interviewee 7, Orthogeriatrics)

Variations in service design were also noted to stem from differences in contextual and regional constraints, including existing structures.

> *So when you look forward into pathways for periprosthetic care, you just have to be cognisant of a) the underlying variation, but b) there is no one size fits all for different ICBs and regions […] because in Region A, there’ll be a heavy arthroplasty centre and so they’re always going to go there and trying to do anything different causes chaos. Region B might be more disparate and so a new hub might make sense, […] we’re terrible in the UK at trying to enforce standardisation on different regions* (Interviewee 10, Orthopaedic and Trauma Surgery)

When rationalising why certain models of care existed, interviewees highlighted local staffing profiles as influencing emerging interdependencies between sites.

> *I think the biggest challenge at the moment is staffing […] when you’re talking about the orthogeriatric input, you can start thinking, well, have you got a consultant, how many whole time equivalent consultants have you got, how frequently are they seeing the patients, are they fully shared care or are they not, so in other words, are you looking after them, doing regular ward rounds, managing them as you would as a medical patient or are you popping in three times a week, seeing anyone that the orthopaedics are worried about and so, the models vary to some extent depending on that.* (Interviewee 2, Orthogeriatrics)

Moreover, geographical distance and established local transportation routes influenced transfer patterns.

> *Hub and spokes work really well if you’ve got a dirty great city with people from one side of the city just going over to the other side of the city for surgery, but if it means going from this side of the range of mountains to that side of the range of mountains, from this valley to that valley […] that’s less attractive to patients so less attractive to government and I’m not sure that hub and spoke developments will happen so quickly […]* (Interviewee 7, Orthogeriatrics)

Transfer of patients across sites, an inherent feature of hub and spoke models, was believed to have both advantages and disadvantages. The advantages were linked to more specialist care, whereas disadvantages were principally related to the longer wait for surgery. This delay had negative implications for both the surgery and postoperative rehabilitation, as POPFF patients who experience delays in receiving surgery are at risk of deconditioning.

> *The biggest factor is really around the deconditioning element of patients. So where there is a transfer it’s not unreasonable that patients might have been waiting 5 days for an operation, […] So patients usually day 1 they’re usually really dizzy when you try to get them up because they’ve been flat on their backs for 5 days, so day 1 is a wipe out […] but also they can be absolutely petrified because they nearly fainted […] So there’s an immediate impact around that deconditioning, fatigue, dizziness, nausea type issues that is different with patients who are transferred in.* (Interviewee 6, Physiotherapy)

### Theme 4: Potential strategies to improve POPFF care

Interviewees made several suggestions as to how the care of patients with POPFF could be improved. As a general strategy, it was suggested that there needed to be greater awareness amongst the clinical community about ‘*the size of the problem and the challenges that this group of patients face’* (Interviewee 6, Physiotherapy). It was felt by some that the body of research on POPFF lagged behind hip fracture. A robust evidence base was seen as a prerequisite to improving care of patients with POPFF so that treatment decisions and the commissioning of new services were evidence based.

> *Now what I’d like to see is to make sure that there’s a decent evidence base for what the most appropriate intervention is and for which patient group and that the decisions that are made are made because the specialist knowledge is there.* (Interviewee 2, Orthogeriatrics)

Interviewees suggested specific areas of POPFF management and care that warranted research including the fracture rate with particular implants; factors affecting care after discharge and long-term rehabilitation; the benefit to the patient of prompt surgery in terms of length of stay and discharge destination; and the overall patient experience of admission due to POPFF.

Development of evidence-based guidance for POPFF service design was recognised by our interviewees as a key strategy for improving care of POPFF patients.

> *I don’t think there is a lot of guidance to be honest on how we should be dealing with it, particularly how we should be structuring our trauma services.* (Interviewee 3, Orthopaedic and Trauma Surgery)

The introduction of financial incentives was another way in which interviewees believed that POPFF care could be improved. Introducing a best practice tariff for POPFF, similar to hip fractures, was suggested by interviewees.

> *I mean what that best practice tariff taught us is that money talks […] if you can operate early, get some money in extra and release a bed earlier then what’s not to like about that? And they’re prepared then to make the changes in staffing or whatever.* (Interviewee 3, Orthopaedic and Trauma Surgery)

While POPFF care is acknowledged to be resource-intensive, interviewees suggested that there were benefits for the patient associated with investment in staff expertise at high-volume centres. Other suggestions for improving POPFF services included taking a holistic approach to the care and management of POPFF patients, increased orthogeriatrician input, and use of the NHFD KPIs to standardise care.

## Discussion

Our qualitative research has highlighted the many mechanisms and nuances through which a multitude of potential service location, service design and institutional factors influence variations for the experiences and outcomes of POPFF patient care. The themes emerging enhance our understanding of the role that contextual factors play, alongside clinical considerations, in shaping quality of care for POPFF patients in current service models and potentially the design of future services.

From our analysis, we have identified four key areas of opportunity relating to service design that warrant consideration when undertaking development of services for patients with.

### 1. Conceptualising POPFF as a sub-specialty

Our interviews highlighted that uncertainty existed over how to conceptualise patients with POPFF. Are they part of the remit of an established field or an emerging specialisation? Are they similar enough to hip fracture patients to benefit from common, established care pathways or do they have distinct needs warranting specialist provision?

The way a problem is conceptualised carries implications for where solutions are sought. It appears that in some hospitals, patients with POPFF follow the same pathway as patients with hip fracture, yet some interviewees clearly identified specific differences from patients with hip fracture, with transfer of patients to an elective or revision hip pathway. In addition, the need for enhanced pain control, blood transfusion, specialist input and timely physiotherapy were highlighted as requirements of effective POPFF care above and beyond standard hip fracture care (4,5). These unique needs of patients with POPFF need to be considered in practice and clinical guidance if aligning care with that for hip fracture is thought to be broadly desirable. The perspectives offered in our study concerning arguments on both sides of this question mirror discourse regarding POPFFs in the contemporary literature. For example, in the background for their study of surgical timing, Murphy et al explain that: “Current guidelines for femoral neck fractures advocate for surgery within 36 to 48 hours … However, the effect of time to surgery is not as clear in periprosthetic hip fractures” (15).

### 2. Implications of patient transfer arising from variable service models

With the exception of some generic literature on the efficiencies of hub and spoke models for concentration of specialist expertise, there is little available evidence for the relative merits of hub and spoke versus self-contained service networks for specific patient groups such as POPFF. Furthermore, more nuanced variations in service models were described amongst our relatively small group of experts, suggesting that there is a gap in current evidence and knowledge concerning how service design should respond to contextual constraints to provide optimal care for this patient group. Although the design of service models for patients with POPFF is not well articulated in the literature, our data shows that they hinge on access to skill, expertise and appropriately equipped units. The consequences of patient transfer delays may be exacerbated for this patient group. A critical issue highlighted by our panel concerns whether the benefits of concentrating expertise in hub units are undermined by the deconditioning that happens when surgery is delayed due to interhospital transfers.

While interviewees tended to describe ‘self-contained’ and ‘hub and spoke’ models, there were also examples of service delivery models that did not fit neatly into these two categories. For example, some sites that might be described as ‘self-contained’ still moved patients to a different site within a larger healthcare Trust. The movement of patients was a source of concern amongst our interviewees as the transfer time was thought to delay surgery and hold significant implications for patient recovery, consistent with the findings of several studies that identified that delayed surgery was associated with an increased duration of post-operative hospital stay (15–17). However, it is currently unknown where the ‘sweet spot’ lies between prompt surgery and the expertise and resource availability that comes with being transferred to a high-volume centre. Furthermore, the existing data is based on quantitative data and is not therefore equipped to account for the reasons why delays may occur (e.g., management of serious medical illness prior to surgery).

More research, and strategies to monitor and evaluate the impact of interhospital patient transfers for POPFF surgery on patient experience and clinical outcomes, is now warranted.

### 3. Integrating multiprofessional support for holistic care

Although our expert panel study sought representation of multiple professional perspectives on POPFF care, a consistent theme and implication of our analysis was the emphasis that experienced professionals placed on integrated, multidisciplinary care for patients with POPFF. Local service configurations may vary in their emphasis in terms of specialty involvement and leadership of different aspects of the POPFF care pathway, but the mutually supportive nature of the functions involved was emphasised consistently during interviews. The success of POPFF management and care depends on more than just effective surgical management, for example. Optimal surgical care will likely be undermined if the patient is unable to take diet or fluids or not supported to mobilise after surgery (18). The potential requirements for extended rehabilitation and multidisciplinary support for patients with POPFF as a distinct group of patients (with more complex fractures requiring specialist treatment, and a high incidence of comorbidity and frailty) should be considered (2). The importance of orthogeriatric input, particularly in the preoperative stages, was emphasised in our data, along with the importance of designing a robust, supported rehabilitation phase into the service model.

### 4. Stimulating future service development

Interviewees suggested greater awareness about the specific needs of patients with POPFF amongst the clinical community is needed if services are to develop and improve. POPFF management and care are under-researched (1), resulting in critical gaps in knowledge to inform service design and good quality guidance. Based on experience in many areas of care (Heart failure, adult asthma, COPD hip fracture), national initiatives that encourage practice developments, such as financial incentives, are likely to be important across the POPFF patient pathway, as are investment in material and human resources (19). Clarity should be sought on the applicability of current hip fracture key performance indicators and associated incentive schemes to POPFF patients as a distinct group.

#### Conclusion and recommendations

This qualitative study has highlighted the many mechanisms and nuances through which a multitude of potential service location, service design and institutional factors influence variations in local service configuration, with subsequent implications for the experiences and outcomes of POPFF care for patients. Understanding the role that contextual factors, alongside clinical considerations, play in shaping the quality of care for POPFF patients in current service models is an important step in enhancing provision for this group through the design of future services. Further research in this area might focus on factors affecting care after discharge and long-term rehabilitation; the benefit to the patient of prompt surgery in terms of length of stay and discharge destination; and the overall patient experience of admission due to POPFF. Research in these areas is warranted to inform such guidance so that it is sensitive to contextual factors driving variation in services.

## Declarations

### Ethics approval and consent to participate

This study was approved by Leeds University School of Psychology Research Ethics Committee (reference: PSCETHS – 722) and was conducted in accordance with the Declaration of Helsinki. Informed consent was obtained from all the participants.

### Availability of data and materials

The datasets generated and/or analysed during the current study are not publicly available to protect participant confidentiality but are available from the corresponding author on reasonable request.

## Data Availability

The minimal data set [and accompanying code, where generated] is available upon request to the authors.

## Acknowledgements

JB is part-supported by the National Institute for Health Research (NIHR) Yorkshire and Humber Patient Safety Research Collaboration. AB’s Unit is an academic unit in the Department of Primary Care and Public Health, within the School of Public Health, Imperial College London. The Unit at Imperial is affiliated with the NIHR Imperial Biomedical Research Centre (BRC) and the NIHR Imperial Patient Safety Research Collaboration. The NIHR Imperial Patient Safety Research Collaboration is a partnership between the Imperial College Healthcare NHS Trust and Imperial College London. The views expressed are those of the author(s) and not necessarily those of the NIHR or the Department of Health and Social Care. MW is affiliated with the National Institute for Health Research Bristol Biomedical Research Centre, University Hospitals Bristol and Weston NHS Foundation Trust and University of Bristol.

